# Tracking multi-site seizure propagation using ictal high frequency activity

**DOI:** 10.1101/2020.05.08.20095323

**Authors:** Steven Tobochnik, Lisa M. Bateman, Cigdem I. Akman, Deepti Anbarasan, Carl W. Bazil, Michelle Bell, Hyunmi Choi, Neil A. Feldstein, Paul F. Kent, Danielle McBrian, Guy M. McKhann, Anil Mendiratta, Alison M. Pack, Tristan T. Sands, Sameer A. Sheth, Shraddha Srinivasan, Catherine A. Schevon

## Abstract

**Objective:** Characterization of progressive multi-site seizure recruitment using high frequency oscillations.

**Methods:** Ictal and interictal high frequency oscillations were identified in a series of 13 patients with 72 seizures recorded by stereotactic depth electrodes, using previously validated methods. Channels with ictal high frequency oscillations were assigned to distinct spatial clusters, and seizure hubs were identified by stereotypically recruited non-overlapping clusters. Clusters were correlated with asynchronous seizure terminations to provide supportive evidence for independent seizure activity at these sites. The spatial overlap of ictal and interictal high frequency oscillations were compared.

**Results:** Ictal high frequency oscillations were detected in 71% of seizures and 10% of implanted contacts, enabling tracking of contiguous and noncontiguous seizure recruitment. Multiple seizure hubs were identified in 54% of cases, including 43% of patients thought preoperatively to have unifocal epilepsy. Noncontiguous recruitment was associated with asynchronous seizure termination (Odds Ratio=10, 95% CI 2.9-41, p<0.001). Interictal high frequency oscillations demonstrated greater spatial overlap with ictal high frequency oscillations in cases with single seizure hubs than in those with multiple hubs (100% vs 66% per patient, p=0.03).

**Significance:** Analysis of ictal high frequency oscillations can serve as a useful adjunctive technique to distinguish contiguous seizure spread from propagation to remote seizure sites. This study demonstrated that multiple seizure hubs were commonly identified by spatial clustering of ictal high frequency oscillations, including in cases that were considered unifocal. The distinction between initially activated and delayed seizure hubs was not evident based on interictal high frequency analysis, but may provide important prognostic information.

Key Points
- Spatial clustering and temporal activation sequences of ictal HFOs were analyzed in a cohort of surgical epilepsy patients.
- Ictal HFOs revealed recruitment of multiple noncontiguous seizure hubs during seizure propagation, even in cases thought to be unifocal.
- The presence of multiple seizure hubs was associated with asynchronous seizure terminations.
- Interictal HFOs showed greater spatial overlap with ictal HFOs in cases with single compared to multiple seizure hubs.

## Introduction

Predictors of post-operative seizure control are needed to help address the limitations of surgical resection and laser ablation in controlling focal drug-resistant epilepsy (DRE).^1-3^ Multiple seizure onset sites are a strong indicator of poor outcome,^4^ however the predictive value of sites that are not active at onset but emerge later in the seizure is uncertain. Few studies have addressed this question, with inconclusive findings.^5-8^

Previously we proposed that seizures are spatially organized into dual zones characterized by distinct neuronal firing patterns: an ictal core, in which synchronized paroxysmal depolarizations are present, and a surrounding penumbra, in which excitatory barrages from the core elicit a strong inhibitory response.^9^ These cannot be reliably differentiated using standard visual EEG interpretation,^9,10^ but can be distinguished by ictal high frequency oscillations (HFOs) at 80-150 Hz, demonstrating persistent phase-locked activity with the EEG, and validated using human microelectrode recordings.^11-13^ Ictal HFOs from clinical electrodes thus serve as a method to track the progressive invasion of synchronized neuronal population firing across the cortex, taking advantage of the strong high gamma signature produced by paroxysmal depolarizing shifts.^14,15^ The order of detection of ictal HFOs relative to seizure onset may modulate the predictive value on surgical outcome,^16^ such that different subcategories of pathological HFOs exist.

We hypothesize that ictal HFOs are organized into noncontiguous spatial clusters, and that these may represent independent seizure hubs, even in cases with a single seizure type and onset zone. Using ictal HFO clustering, activation sequences, and seizure termination patterns, we characterize the dynamic spatiotemporal properties of seizure recruitment, and define patient-specific seizure hubs in consecutive patients with focal DRE implanted with depth electrodes. We also compare the spatial overlap between ictal and interictal HFOs to evaluate whether interictal HFOs are sensitive to temporal sequences of seizure recruitment. This study provides a framework for implementing ictal HFO analysis into the surgical evaluation of poorly localized and multifocal epilepsy.

## Methods

### Patient Description

Thirteen consecutive patients at Columbia University Medical Center (CUMC) undergoing surgical evaluation using implanted depth electrodes (PMT Corporation, Chanhassan, MN) between 2014 and 2016 were included. All clinical seizures were included for each patient with two exceptions. In one patient who required electrode revision, only post-revision seizures were included. Another patient had numerous focal aware seizures, of which nine were randomly selected for analysis. All protocols for Human Subjects Research were approved by the CUMC Institutional Review Board with informed consent.

### EEG Analysis

Intracranial EEG was recorded using the XLTek EEG system (Natus Medical Incorporated, Pleasanton, CA), sampled at 500 Hz, with an epidurally placed mini-depth electrode reference. Seizure onset zone (SOZ) was defined as the channel(s) with the first electrographic ictal activity by visual inspection. Epilepsy syndromes were considered unifocal if a single SOZ was present across all seizures. As a proxy for the potential of a given site to generate seizure activity independently, we assessed correlation with asynchronous seizure termination. Asynchronous termination was identified by a difference in termination of the ictal rhythm in multiple channels of at least 1s, or a difference of at least 500ms if the rhythm across channels had clearly different morphology and frequency at seizure termination. Channels were then accordingly divided into non-overlapping termination groups. All EEG data were reviewed independently by at least two of the authors (ST, LMB, CAS), and disagreements were resolved by consensus.

### Ictal HFO Analysis

Ictal HFOs were visually identified from filtered EEG data (FIR, order 90, bandpass 80-150 Hz) displayed using Persyst Insight (Persyst Development Corp, Solana Beach, CA), and validated by comparison to the MATLAB FIR1 filter with the same parameters (Mathworks, Natick, MA), similar to those used previously.^11,16^ Ictal HFOs were defined as previously described, with morphology including at least four consecutive peaks clearly standing out from the background,^17^ and required to demonstrate delayed activation after seizure onset with repetitive bursting activity for at least 10 seconds, and visual correlation with the low-frequency EEG rhythm (Fig. 1B). This was intended to mirror the quantitative phase-locked high gamma method previously developed to track the path of seizure recruitment,^11,16^ and allow for simple translation to clinical use. HFOs were identified blinded to imaging coregistration and electrode location. Ictal HFOs attributable to artifact (e.g. an identical pattern across multiple channels) were excluded. Additionally, channels were excluded if they demonstrated significant interictal artifact, or questionable high frequency signal that was non-sustained. In focal to bilateral tonic-clonic (FBTC) seizures, HFO analysis was conducted up to the time of clinical transition to bilateral tonic-clonic activity with associated generalized ictal EEG rhythm, as identified by video-EEG inspection. For all other seizure types, HFO analysis was performed from electrographic onset to termination.

**Figure 1.**
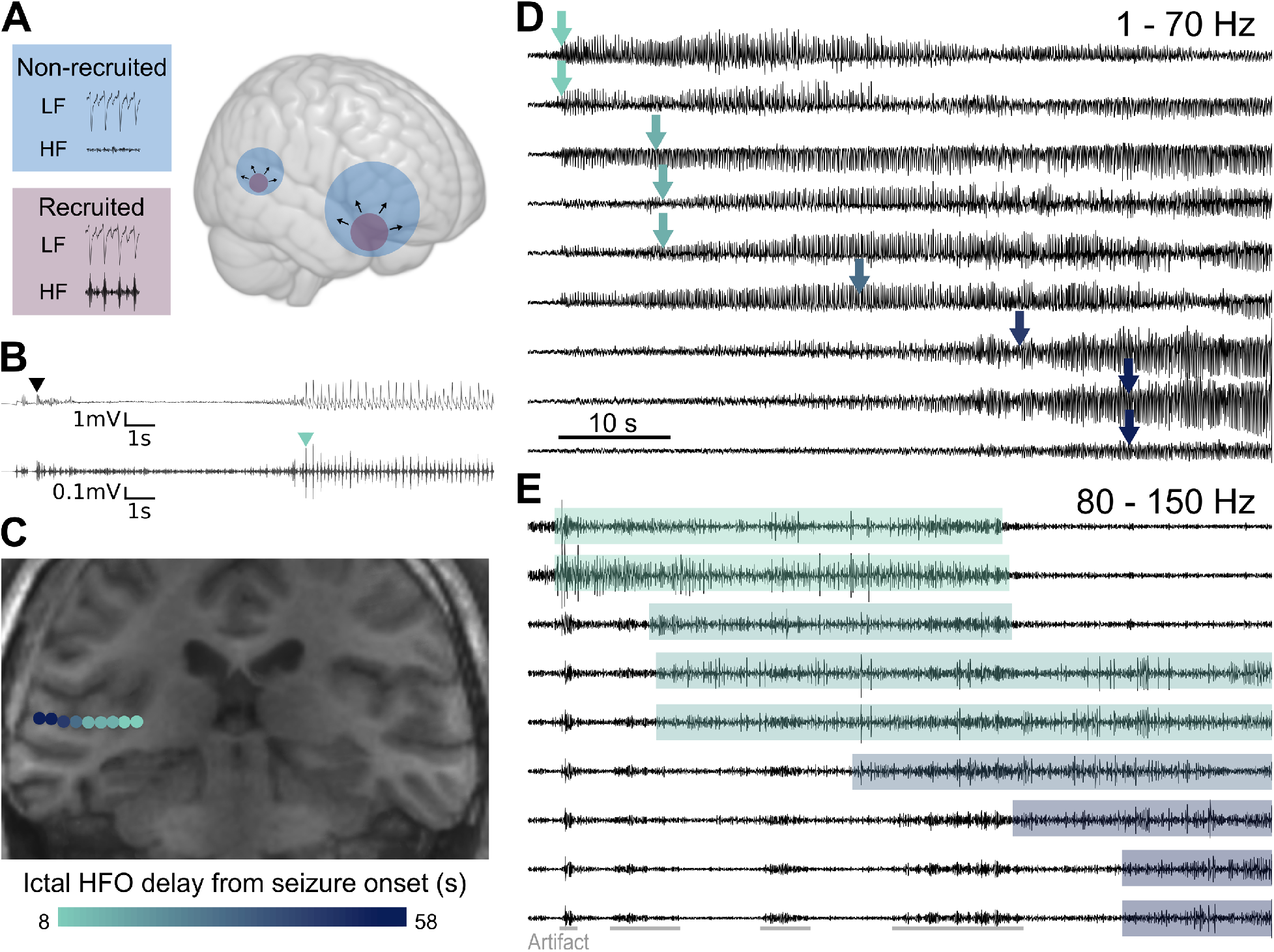
Local propagation within a seizure hub. A. Schematic model of the spatial structure of seizures, depicting multiple seizure hubs with expanding recruited territory identified by ictal HFO activity phase-locked to the ictal EEG rhythm, surrounded by non-recruited regions involving an ictal EEG rhythm without ictal HFOs. B. Example of visual analysis of delayed ictal HFO activation. Seizure onset in standard EEG (top trace, black arrowhead) precedes the initiation of repetitive HFO bursts (bottom trace, green arrowhead) that correlate with low frequency ictal EEG discharges. C. Localization and sequential activation of ictal HFOs along the depth array in the superior temporal gyrus, indicating a slow radial expansion of seizure recruitment. D. Standard EEG reveals early activity in the majority of channels. Arrows indicate timing of ictal HFO activation. E. Ictal HFOs (80-150 Hz) demonstrate ordered recruitment that is not well appreciated in (D). LF, low frequency; HF high frequency.

### Interictal HFO Analysis

Intracranial EEG segments with durations of 10 minutes of NREM sleep and free of extracerebral artifact were visually identified for each patient. Segments were chosen at least 24 hours after implantation to minimize anesthetic effect and four hours from the most recent seizure if possible. As the majority of interictal HFOs (>99%) were detected in association with epileptiform discharges, we aimed to minimize bias due to variable representation of multifocal discharge populations in the selected EEG segments. Multifocal discharge populations were defined as epileptiform discharges with non-overlapping fields in different anatomic structures. Each discharge population was visually identified, and if necessary, an additional extended segment beyond 10 minutes was selected to include 20 representative discharges from each region showing independent discharge populations. Extended segments were needed in three cases.

Automated interictal HFO detection in the ripple band was performed using the short time energy (RMS) method in RIPPLELAB.^18^ A minimum cutoff of eight oscillations was used to decrease false positive HFO identification, as previously described.^19^ Otherwise there were no changes to the default parameters. All automated HFO detections were reviewed using simultaneous views of the unfiltered event, 80-250 Hz filtered event, and spectral density plots for the given channel. The interictal record was also visually reviewed for the presence of an epileptiform discharge associated with each interictal HFO identified in the same or any other channel. Artifacts were excluded using similar criteria as for the ictal analysis. After visual inspection of detections and exclusion of false-positives, HFO rates were determined for each initial 10-minute segment, as well as for the extended segments. An HFO asymmetry index was used to evaluate the association between interictal HFOs and the SOZ, calculated per patient by taking the difference between interictal HFO rates inside and outside of the SOZ, then dividing by the sum of rates.^20,21^ This index ranges from −1 to +1, such that values closer to +1 indicate higher HFO rates inside the SOZ, whereas values closer to −1 indicate higher rates outside the SOZ.

### Cluster Analysis

Electrode localization was performed by coregistration of the pre-implant T1 volumetric brain MRI with the post-implant volumetric head CT using FSL (FMRIB Software Library, FMRIB, Oxford, UK). After coregistration, contacts were manually localized individually. Distinct clusters were defined per seizure as sites with ictal HFOs in different anatomic structures and separated by at least 2 cm, a distance chosen to limit the effect of local hypersynchronous activity.^22^ For each cluster, an index contact was identified as the channel with earliest HFO onset within that cluster or with the highest amplitude if there were multiple channels with simultaneous onset. Cluster distances were determined by the Euclidean distances between index contacts. Propagation between clusters was visually defined by sequentially activated index contacts. Simultaneous cluster activation was defined by <200 ms between activation of index contacts. Seizure hubs were defined for each patient by aggregating all ictal HFO sites in analyzed seizures, and identifying non-overlapping regions using the same criteria as for ictal HFO clusters.

### Statistical Analysis

Comparison between continuous data were evaluated using the Mann-Whitney U-test (for two samples) or Kruskal-Wallis H-test (for greater than two samples). Associations between categorical data were evaluated using Fisher’s exact test. Correlation was calculating using the Spearman correlation coefficient. All statistical tests were two-sided with a significance threshold of 0.05. All analyses were performed using R 3.5.3 (R Foundation, Vienna, Austria).

### Data Availability

Data will be shared with other investigators upon request and subject to limitations imposed by the CUMC Institutional Review Board, Information Technology policies, and federal medical data privacy laws.

## Results

Seventy-two seizures from 13 patients (mean 5.5 seizures per patient, range 2-20) were analyzed. These included 45 focal impaired awareness seizures (FIAS), 15 focal aware seizures (FAS), and 12 FBTC seizures. Presurgical characteristics are shown in Table 1. Ictal HFOs were identified in 12/13 (92%) patients and 51/72 (71%) seizures. Among seizures without ictal HFOs (16 FIAS, two FAS, three FBTC), there were 15 brief seizures with durations less than 45 seconds (13 FIAS, two FAS), one FIAS with a limited field of ictal discharges, and five limited by excessive artifact (two FIAS, three FBTC).

**Table 1.** Patient demographics and presurgical characteristics.

| ID | SEX | AGE | SEIZURE TYPE | NUMBER OF SEIZURES | MRI | PET | PRE-OP SUSPECTED MULTIFOCAL |
| --- | --- | --- | --- | --- | --- | --- | --- |
| 1 | F | 53 | FIA | 3 | Nonlesional | R mesial temporal | No |
| 2 | F | 50 | FIA; FBTC | 3 | L MTS; multiple bilateral cavernomas | L mesial and temporal pole | Yes |
| 3 | F | 30 | FA | 5 | R choroidal fissure cyst | R mesial and temporal pole | No |
| 4 | M | 25 | FIA | 4 | L frontal cortical encephalomalacia | L frontal cortical and basal ganglia/thalamus | No |
| 5 | M | 23 | FIA; FBTC | 4 | L choroidal fissure cyst | Bilateral anterior temporal | Yes |
| 6 | M | 30 | FIA | 4 | Nonlesional | Bilateral temporal | Yes |
| 7 | F | 52 | FIA | 2 | L temporoparietal signal and possible cortical thickening; R aneurysm clipping | L mesial/anterior temporal | Yes |
| 8 | F | 29 | FBTC | 2 | Punctate L medial frontal and R posterior temporal susceptibility | R>L temporal | No |
| 9 | F | 31 | FIA | 4 | L temporo-parieto-occipital band heterotopia; possible L hippocampal atrophy; dandy-walker variant | L mesial and temporal pole | Yes |
| 10 | F | 29 | FA | 9 | L MTS | L anterior hippocampus | No |
| 11 | F | 55 | FIA | 20 | Possible R frontal subependymal signal abnormality | R anteromedial temporal | No |
| 12 | M | 25 | FA; FBTC | 3 | Bilateral posterior periventricular heterotopias | R temporal | Yes |
| 13 | M | 19 | FIA; FBTC | 9 | Postsurgical R temporal encephalomalacia | R hemispheric | No |

FA, focal aware; FIA, focal impaired awareness; FBTC, focal to bilateral tonic-clonic; MTS, mesial temporal sclerosis

### Ictal HFO Spatial Clustering

Ictal HFOs were detected in an average of 10% (range 0-20%) of the total implanted contacts per patient. Ictal HFOs were recorded exclusively from gray matter structures, with the exception of one case in which two contacts located extra-axially in CSF adjacent to epileptogenic cortex consistently recorded ictal HFOs. A total of 88 ictal HFO clusters were identified from the 51 seizures, with a median of 4.5 (range 1-21) electrode contacts per cluster. As detailed in Table 2, ictal HFOs were confined to a single cluster in 25 seizures from 10 patients, of which 21 were in mesial temporal regions and four were neocortical (three temporal, one frontal). Among the five patients with bilateral implantation, ictal HFO clusters were found bilaterally in five seizures from two patients, with all involving both mesial temporal and lateral temporal sites.

**Table 2.** Ictal HFO localization and surgical characteristics.

| ID | SEEG SOZ | ICTAL HFO CLUSTER LOCALIZATION | SEIZURE HUBS | SURGERY | ENGEL OUTCOME | FOLLOW-UP (MO) |
| --- | --- | --- | --- | --- | --- | --- |
| 1 | R MT | R amygdala, hippocampus | Single | Inferior hippocampus + parahippocampal gyrus laser ablation | IIA | 49 |
| 2 | L MT | L hippocampus | Single | Hippocampal head/body + amygdala laser ablation | ID | 29 |
| 3 | R MT | R hippocampus | Single | Hippocampal head/body laser ablation | IB | 18 |
| 4 | L frontal perilesional (clinical); L MT (subclinical) | L superior frontal gyrus; L inferior parietal lobule; L posterior cingulate; L hippocampus | Multiple | NA | NA | – |
| 5 | Bilateral MT | L hippocampus; R hippocampus; R inferior temporal gyrus | Multiple | NA | NA | – |
| 6 | Bilateral MT; L orbitofrontal | L entorhinal cortex; L parahippocampal gyrus; R middle temporal gyrus | Multiple | NA | NA | – |
| 7 | L MT | L hippocampus | Single | Hippocampal head/body + amygdala laser ablation | IIB | 34 |
| 8 | L frontal neocortical | NA | NA | Medial frontal cortical lesionectomy | IA | 18 |
| 9 | L temporoparietal neocortical | L parietal heterotopia; L temporal heterotopia | Multiple | Temporoparietal corticectomy | IIIA | 25 |
| 10 | L MT | L hippocampus; L parahippocampal gyrus | Multiple | Hippocampal head/body + amygdala laser ablation | ID | 24 |
| 11 | R temporal neocortical | R hippocampus | Single | Extended temporal lobectomy | IB | 6* |
| 12 | R posterior perilesional; R MT | R hippocampus; R parahippocampal gyrus; R middle temporal gyrus; R superior temporal gyrus | Multiple | Hippocampal head/body + medial occipital laser ablation | IVA | 22 |
| 13 | R temporal neocortical | R superior temporal gyrus; R inferior parietal lobule; R fusiform gyrus | Multiple | Anterior temporal lobectomy + superior temporal gyrus and posterior middle temporal gyrus resection | ID | 21 |

\* Long-term follow-up data not available
SOZ, seizure onset zone; MT, mesial temporal

The extent of ictal HFO spatial clustering was evaluated against clinical seizure type. There was no difference in the number of ictal HFO clusters or the number of contacts per cluster between seizures with or without impaired awareness (n=51 seizures, mean 1.8 vs 1.5 clusters per seizure, U=269, p=0.61; mean 5.7 vs 6.5 contacts per cluster per seizure, U=218, p=0.53). Analysis of only the first four seizures per patient had similar results (data not shown). Five patients had FBTC seizures, with ictal HFOs recorded in nine seizures from three patients. All these seizures had multiple clusters, with an increased number of clusters compared to FIAS without generalization (n=38 seizures, mean 2.6 vs 1.6 clusters per seizure, U=214, p=0.002).

### Ictal HFO Activation Sequences

To evaluate seizure propagation and the implied expansion of recruited territory, we assessed activation sequences of ictal HFO activity. Sequential activation of ictal HFO channels was observed within single clusters (Fig. 1), as well as between distinct clusters (Fig. 2). In six seizures from three patients, there was simultaneous activation of clusters, indicating nearly instantaneous recruitment of sites separated by Euclidean distances ranging from 2.0-4.7 cm. Excluding these simultaneously activated clusters, the mean delay between the first and second activated clusters was 37.2 sec (6-182 sec), and between second and third clusters 35.0 sec (7-68 sec). From these figures, a mean propagation speed of 2.15 mm/sec (range 0.19-6.0 mm/sec) was calculated between the first and second non-simultaneously activated clusters (Fig. 3A).

**Figure 2.**
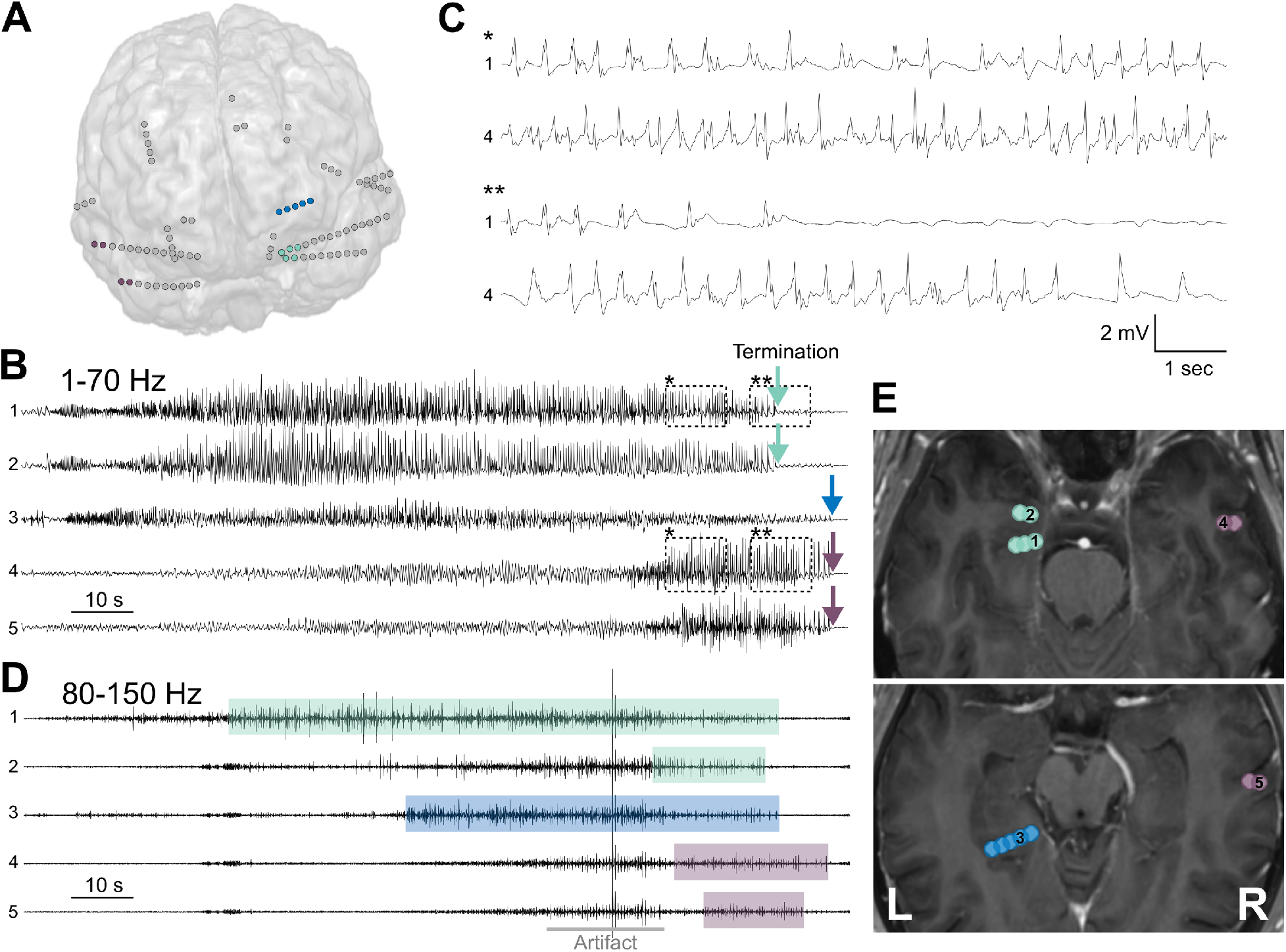
Remote propagation between seizure hubs. A. Ictal HFO activity identified in three distinct clusters of contacts (color-coded), representing a minority of the contacts that recorded ictal EEG rhythms (gray). B. Ictal EEG (1-70 Hz) demonstrating asynchronous terminations (arrows). C. Ictal EEG traces from contralateral sites show asynchronous rhythms (*) leading into asynchronous terminations (**), suggestive of independent seizure foci. D. High frequency (80-150 Hz) activity reveals an ordered pattern of recruitment across ictal HFO clusters. Artifact may appear similar to HFO activity, particularly in narrow time-base view. Non-sustained HFOs are seen in Channel 2 prior to development of sustained activity distinct from artifact. E. Ictal HFO localization reveals propagation from entorhinal cortex to parahippocampal gyrus to contralateral temporal neocortex.

**Figure 3.**
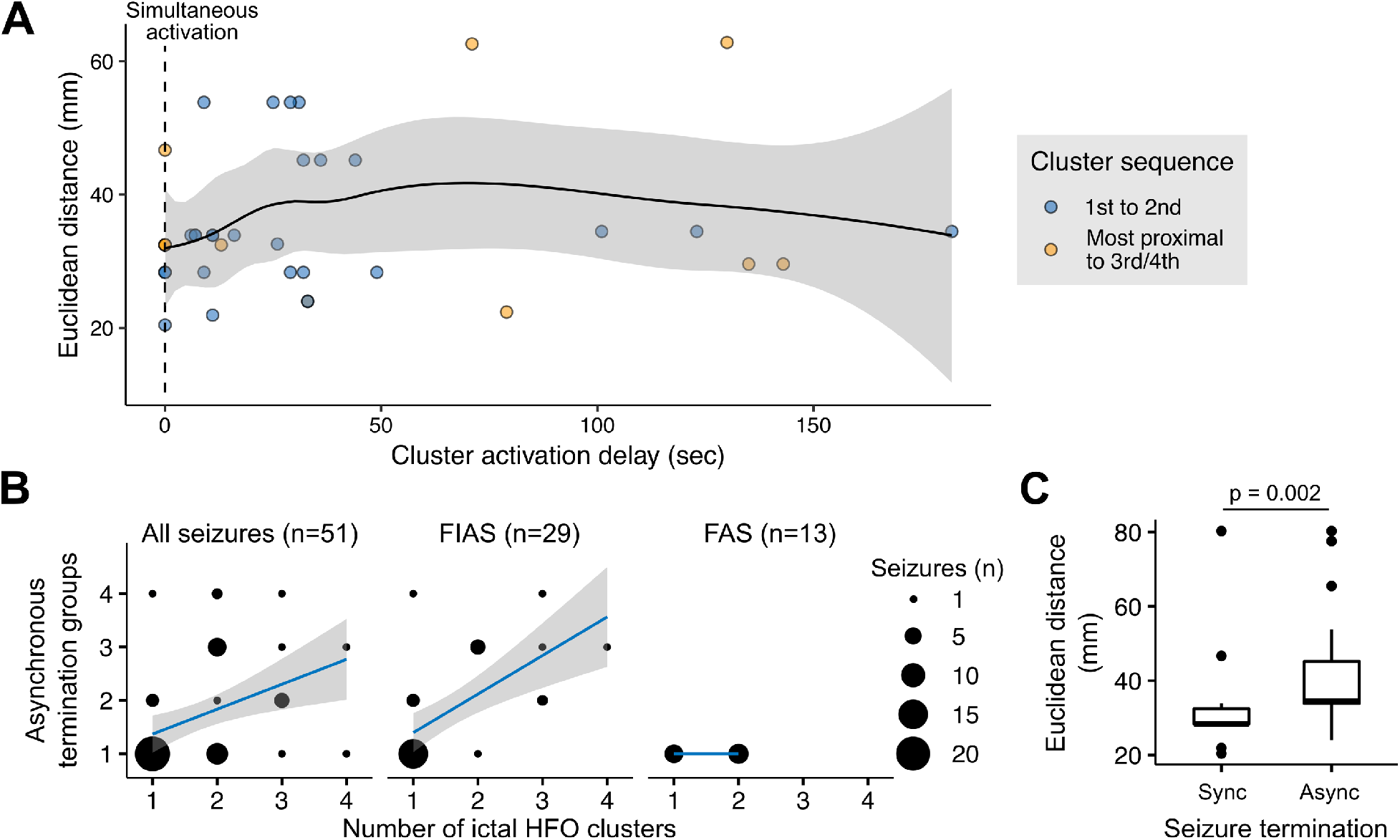
Inter-cluster distances and termination synchrony. A. Inter-cluster delays plotted against Euclidean distances between the first-to-second activated clusters (blue) and third or fourth activated clusters from the most proximally located earlier cluster (yellow). There is evidence of a nonlinear association involving both rapid and slow propagation between clusters, including nearly simultaneous activation of distinct clusters. Gray shaded area represents the 95% confidence interval. B. The number of ictal HFO clusters per seizure correlates with the number of groups of channels terminating asynchronously. When stratified by seizure type, this association is strongest in focal impaired awareness seizures (FIAS), and is not seen at all in focal aware seizures (FAS) due to absence of asynchronous terminations. Gray shaded area represents the 95% confidence interval. C. There is a greater distance between sequentially activated ictal HFO clusters in seizures terminating asynchronously compared to those terminating synchronously.

### Ictal HFO Clusters and Asynchronous Termination

To establish that subsequent ictal HFO clusters represent independent sites, we assessed seizure termination patterns of cluster sites. Asynchronous termination was observed in 22 seizures, including 20/51 (39%) seizures with ictal HFOs and 2/21 (9.5%) without ictal HFOs. Asynchronous termination was more commonly observed in seizures with multiple ictal HFO clusters compared to seizures with single or no clusters (n=72, Odds Ratio=10, 95% CI 2.9-41, p<0.001). The number of ictal HFO clusters per seizure correlated positively with the number of channel groups terminating asynchronously (n=51, r_s_=0.48, p<0.001). When stratified by seizure type, this correlation became stronger for FIAS (n=29, r_s_=0.69, p<0.001) and was no longer observed for FAS due to complete absence of asynchronous termination in this group (Fig. 3B). Pairs of contacts within the same cluster were more likely to share the same termination group than pairs from different clusters (n=75, mean proportion per seizure 1.0 vs 0.54, U=973, p<0.001). In 13 seizures from five patients, the sequential order of seizure termination followed the order of cluster activation, such that a cluster activated after another also terminated afterwards. In contrast, there was only one seizure in which a cluster activated after another showed earlier seizure termination. In 13 instances, there were no ictal HFO clusters corresponding to the termination group. Among seizures with multiple clusters, distances between sequentially activated clusters were greater in seizures with asynchronous termination compared to those with synchronous termination (n=37, mean 42.6 vs 33.6 mm, U=59.5, p=0.002, Fig. 3C).

### Patient-level Seizure Hubs

The contacts comprising ictal HFO clusters showed consistent seizure-to-seizure activation, indicative of stereotyped seizure hubs per patient. There were 26 seizure hubs of unique non-overlapping ictal HFO clusters. The mean and median proportion of seizures in which ictal HFO contacts were activated when the corresponding seizure hub was recruited were 0.83 and 1.0, respectively. The index contact, representing the channel activated first within a cluster, remained invariant in 25/26 (96%) of seizure hubs.

There were a median of 2 (range 0-4) seizure hubs per patient. Preoperatively, 6/13 (46%) cases were suspected to have multifocal onsets (including two cases of bitemporal epilepsy), and multiple seizure hubs were found in 7/13 (54%) cases. However, there was limited overlap between these groups, with multiple seizure hubs in 3/7 (43%) cases suspected pre-operatively to have unifocal epilepsy and in 4/6 (67%) cases suspected to have multifocal involvement, including both bitemporal cases (Table 2).

### Ictal and Interictal HFO Overlap

Interictal HFOs at 80-250 Hz were identified in 9/13 (69%) cases. The mean interictal HFO rate per patient was 30 (1-69) per 10 minutes, with a mean individual channel rate of 4.2 (1-31) per 10 minutes. There were 70 unique channels (4.3% of total implanted) demonstrating interictal HFOs, with 5 channels identified solely from extended segments used to ensure adequate sampling of all discharge populations. Interictal HFOs were correlated with the SOZ, as indicated by a positively skewed HFO rate asymmetry index (mean 0.23, 95% CI -0.19-0.65).

We next assessed the spatial correlation of interictal with ictal HFOs (Fig. 4). There was ictal and interictal HFO overlap in 71% of all interictal and 30% of all ictal HFO contacts. In patients with both ictal and interictal HFOs, interictal HFOs were observed within 8/19 (42%) of seizure hubs. Among the 20 channels with interictal HFOs located outside of seizure hubs, 7 (35%) were from immediately adjacent contacts. The proportion of interictal HFO channels within seizure hubs was dependent on the number of hubs per patient (Fig. 4A). In cases with single seizure hubs, 100% of interictal HFO channels were found within the hub, compared to cases with multiple seizure hubs, in which an average 66% of interictal HFO channels per patient were found within hubs (n=9, U=0, p=0.03). There was no relationship between the order of seizure hub activation and interictal HFO overlap (n=17, H_3_=2.2, p=0.53) or rate (n=17, H_3_=1.8, p=0.61), nor in interictal HFO rates between the earliest activated seizure hub and all later activated hubs combined (n=65, 16.0 vs 11.4 interictal HFOs per minute, U=617, p=0.22).

**Figure 4.**
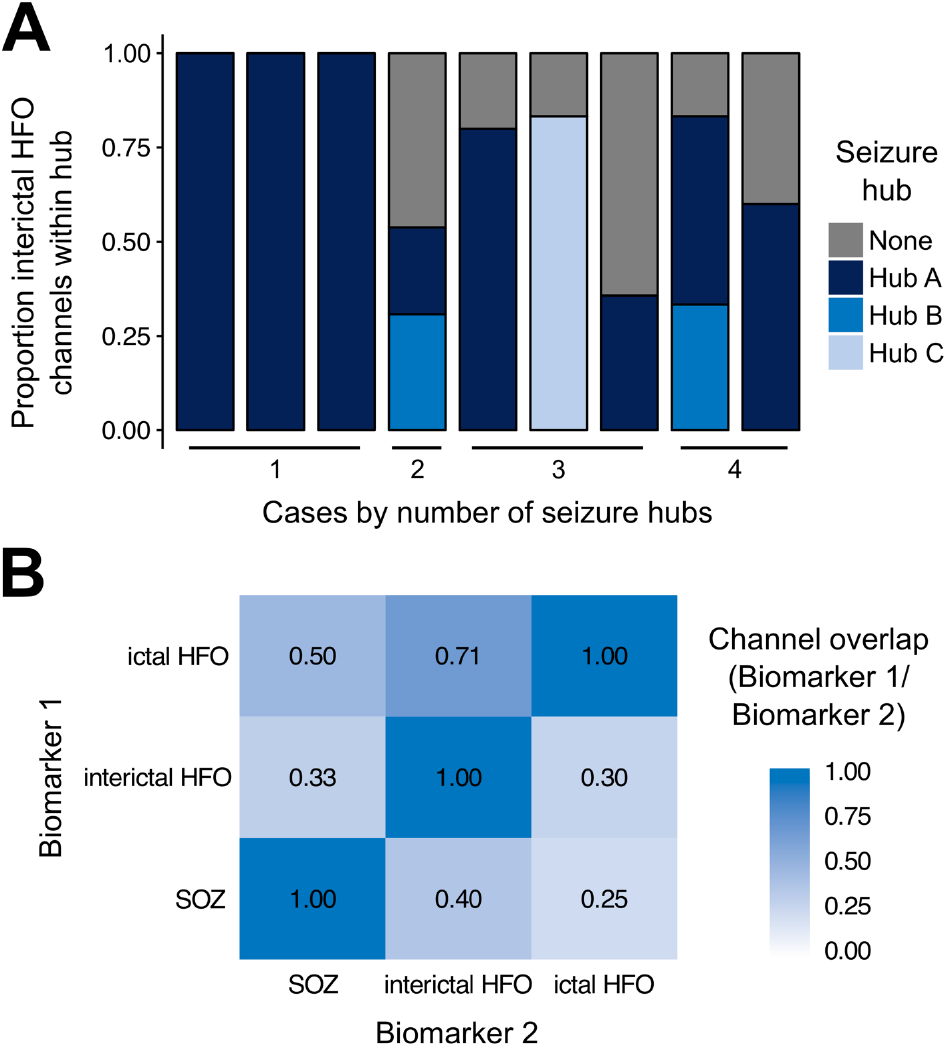
Association between ictal HFOs, interictal HFOs, and SOZ. A. In patients with a single seizure hub, all interictal HFOs channels localize to that hub. Across patients with multiple seizure hubs, interictal HFOs are found in 8/19 (42%) of hubs, and 29% of interictal HFOs are located outside of seizure hubs. B. Proportion of spatial channel overlap between ictal HFO, interictal HFO, and SOZ biomarkers.

### Surgical Outcomes

All patients with single seizure hubs underwent surgery and had Engel Class I/II outcomes (Table 2). The two patients with Engel Class III/IV outcomes had multiple seizure hubs and interictal HFOs outside the SOZ. Three of the five remaining patients with multiple hubs were not considered candidates for resection or laser ablation. Good outcomes were seen in 5/5 (100%) patients in whom all seizure hubs were entirely removed, compared to 2/4 (50%) with at least one hub not entirely removed.

## Discussion

In this series of patients with implanted depth electrodes, we investigated the spatial clustering and temporal activation sequence of ictal HFOs, their association with asynchronous seizure termination, and their relationship with interictal HFOs. We found that even in patients with unifocal epilepsy, ictal HFOs revealed recruitment of multiple noncontiguous seizure hubs during seizure propagation, which were associated with asynchronous seizure terminations. In addition, patients with multiple seizure hubs were more likely to have widely distributed interictal HFOs. These delayed-onset seizure hubs are generally not considered in standard surgical decision-making, and may explain incomplete post-operative seizure control in certain cases.

Ictal HFOs occurring in the form described here, with sustained bursting correlated to the low-frequency EEG rhythm, have been previously validated as a biomarker of synchronized neuronal population firing using human microelectrode data,^11^ and in a subsequent surgical outcome study.^16^ It is important to note that this definition differs from that used in earlier ictal HFO studies,^17,23-27^ which would have resulted in inclusion of additional high frequency events, such as non-sustained gamma bursts at the ictal-interictal transition. Apart from the rare instances in which HFOs were identified from extra-axial contacts immediately adjacent to cortex, HFOs were exclusively found in contacts positioned in gray matter structures. This suggests that non-physiologic artifactual effects were not prominent. We found frequent instances (as shown in Fig. 1-2) of a characteristic ictal EEG rhythm without accompanying HFO activity. This behavior is predicted by our prior work distinguishing the recruited ictal core from the surrounding penumbra with disorganized multiunit firing and preserved inhibition,^9^ which may show high-amplitude EEG signals but is less likely to generate HFO signals.^11,14^ Over half of the cases here were found to have multiple distinct seizure hubs based on ictal HFO activity, confirming preoperative suspicions of multifocality in 67% of cases, as well as identifying multiple hubs in 43% of cases presumed to be unifocal. This is likely to be true of many focal epilepsies,^4^ and the use of the method described here may facilitate identification of independent seizure foci more rapidly than other currently available clinical methods.^28^

By analyzing the spatiotemporal properties of ictal HFOs, we found evidence for dual modes of propagation: slow contiguous spread and rapid noncontiguous spread. In the example shown in Figure 1, in which slow contiguous spread through a neocortical gyrus was captured by a single electrode, the recruitment speed (0.57 mm/s) aligns with the propagation speeds established in animal models employing calcium imaging,^29^ as well as propagation speeds calculated from direct measurements of the ictal wavefront recorded in humans.^9^ In contrast, cases in which ictal HFOs appeared nearly instantaneously between sites separated by up to 4.7 cm indicate that contiguous spread through intervening brain is highly unlikely (particularly since spread would follow a longer path than the lower-bound Euclidean distances). This situation is distinct from visual EEG observations of simultaneous fields extending to remote sites, which can occur via synaptic current distribution without necessarily reflecting seizure recruitment at each site.^30^ Instead, simultaneous recruitment at noncontiguous sites must proceed by a different mechanism. One possibility is impaired local inhibition at the secondary site permitting near-instantaneous "jumps" as demonstrated in an animal model using focal bicuculline microinjections at sites remote to 4-aminopyridine seizure foci.^31^ Seizure emergence may therefore be dependent on the state of local inhibition rather than connectivity between sites. The progressive recruitment of multiple seizure hubs observed in this study would be anticipated as a consequence of focally impaired inhibition at vulnerable sites, although direct evidence is lacking. Furthermore, interactions between these hubs and the resulting multiscale EEG dynamics may underlie observations of epileptic networks.^32,33^

The consistent localization, activation sequence, and channel composition of ictal HFO clusters indicate that these hubs are stereotyped from seizure to seizure. However, further investigation is needed to define how these seizure hubs interact with each other and to distinguish which sites contribute to the epileptogenic zone. While resection of early appearing ictal HFOs was a stronger predictor of postoperative seizure freedom than later appearing HFOs in prior work,^16^ it is uncertain whether later appearing HFOs arising from local versus remote seizure spread has prognostic significance. A larger study is required to determine whether the presence of delayed seizure hubs are negative outcome predictors, and whether intervention at these sites may improve outcomes.

Intracranial EEG is inherently limited by sparse spatial sampling, and the apparent SOZ may not be the true site of seizure initiation, as is generally the case for intracranial recordings. Instead, the site of seizure initiation is inferred by considering the timing and location of the electrographic onset in the context of other clinical data. Sparse sampling is a limitation shared by all studies using intracranial electrodes, including prior analyses of HFOs and network models. However, in contrast to prior ictal HFO studies,^17,23-27^ the method we describe here may help to overcome some of these sampling issues. Rather than utilizing HFOs as context-free, equally-weighted biomarkers, we propose that they be used as an adjunct to clinical EEG interpretation, by tracking the path of recruited territory as the seizure advances from a possibly unsampled origin through sampled brain sites. The association between ictal HFO clustering and termination asynchrony, in which those ictal HFO channels in different clusters and separated by greater distance were more likely to terminate asynchronously, may provide a method of detecting undersampling. For example, a group of channels terminating synchronously without ictal HFOs, as seen in a minority of cases here, may imply synaptic spread to these channels from an unsampled seizure hub. The absence of ictal HFOs entirely or appearing only late in the seizure may also suggest recruitment in an unsampled region.

Compared to interictal HFOs, which have also generated significant interest as a biomarker of epileptogenicity,^20,34-39^ ictal HFOs provide necessary temporal information to identify areas recruited earlier versus later in the seizure.^16,17,25,26^ Ictal and interictal HFOs have been previously detected in overlapping channels up to the first five seconds after seizure onset,^40^ but did not characterize later propagation or involvement of multiple sites. While we found concordant localization between ictal and interictal HFOs in cases with a single seizure hub, interictal HFOs in cases with multiple hubs failed to identify the majority of these, while also appearing outside of ictal HFO clusters. Additionally, interictal HFO rates did not distinguish between initially activated versus delayed seizure hubs. These results may help explain recent findings that interictal HFOs are insufficient predictors of epileptogenic zones at the individual patient level.^21,41^ Although interictal HFOs did not provide temporal information about seizure recruitment, we found evidence for spatial clustering together with ictal HFOs. Therefore, since interictal HFOs may have value in the absence of seizures, consideration of both biomarkers together may improve detection of the epileptogenic zone. Interictal HFO rates in this study were lower than in some prior studies,^20,39^ and within the ranges reported by others,^21,36^ likely reflecting differences in the individual parameters and methods used. A conservative approach to interictal HFO measurement was particularly important for this study to avoid false positives at the expense of reduced sensitivity. However, this analysis was confined to ripple band HFOs, and therefore was potentially limited by inability to assess fast ripples, which have a higher reported specificity for epileptogenicity.^36,39^

Due to spatial sampling limitations, it is unlikely that all possible sites generating ictal HFOs were identified. Additionally, the criteria for 2 cm separation distance between clusters may have defined adjacent clusters as distinct, when in fact they were part of the same seizure hub. The criteria requiring clusters to be in distinct anatomic structures was used to minimize this possibility. Analysis of surgical outcomes is limited by the small number of patients, however the purpose of this study was to define a methodology that can be translated directly to clinical use, and to support a larger study addressing the predictive value of ictal HFO analysis on surgical outcomes.

In summary, ictal HFO analysis can track seizure recruitment within and across multiple distinct seizure hubs. We speculate that focal seizures spread broadly not only by synaptic distribution of excitatory currents but also by seeding new hubs that amplify seizure distribution and create the appearance of large-scale coordinated network behavior. A more complete understanding of this process at the level of both EEG recordings and cellular activity is needed to inform the clinical approach in multifocal DRE, particularly in the context of increasingly targeted surgical approaches.

## Acknowledgements

We would like to thank the patients and families, whose contribution and participation are deeply appreciated. We would also like to thank members of the Columbia University Comprehensive Epilepsy Center and the Epilepsy Fellows who contributed to patient care. This work was supported by NIH/NINDS R01-NS084142 (CAS) and R01-NS095368 (CAS).

